# The roles of unrecognized monkeypox cases, contact isolation and vaccination in determining epidemic size in Belgium. A modelling study

**DOI:** 10.1101/2022.07.28.22278048

**Authors:** Christophe Van Dijck, Niel Hens, Chris Kenyon, Achilleas Tsoumanis

## Abstract

We used a network model to simulate a monkeypox epidemic among men who have sex with men. Our findings suggest that unrecognized infections have an important impact on the epidemic, and that vaccination of individuals at highest risk of infection reduces epidemic size more than post-exposure vaccination of sexual partners.

## Background

Monkeypox is a viral zoonosis whose spread was, until recently, almost exclusively limited to Central and West Africa. Since May 2022, over 15,000 cases of monkeypox have been confirmed from every continent excluding Antarctica (https://ourworldindata.org/monkeypox, 24 July 2022). In this multi-country outbreak, the number of cases resulting from human-to-human transmission is much higher than ever reported, and unlike the outbreaks in Africa, many cases bore several hallmarks of sexual transmission. Most cases were young men and where this information was available, typically men who have sex with men (MSM) with high rates of partner change (termed higher risk-, or HR-MSM). [1,2] Furthermore, monkeypox was frequently linked to sexual encounters and presented with localized anogenital lesions compared to the generalized skin lesions typically associated with monkeypox. [1,2] We and others have noted that a sizeable proportion of cases report few, atypical, or absent symptoms. [3] This could have an important impact on transmission of the monkeypox virus. Public health recommendations to contain the epidemic include isolation of cases, requesting close contacts to abstain from sex and pre- or post-exposure (ring) vaccination of individuals at high risk of infection with a smallpox vaccine. [4–6]

Previous modelling studies have estimated that monkeypox has epidemic potential in the general population, but that such epidemics can generally be contained by case isolation, contact tracing and/or ring vaccination. [7–10] These efforts have, thus far, been insufficient to contain the epidemic. [8]

In this manuscript we evaluate the impact of undiagnosed infections on a sexually associated monkeypox outbreak in an MSM sexual network, and we test the hypothesis that contact tracing or vaccination reduce the epidemic. We do this using a network-based model, parameterized with Belgian MSM behavioral data.

## Methods

### Network model

Building on a previously published separable temporal exponential family random graph model of a Belgian MSM population, [11] we added a population of HR-MSM which was parameterized with data from the cohort of HR-MSM that was included in the Preventing Resistance in Gonorrhea (PReGo) study in Belgium [12]. The model was refined to include main and casual partnerships among low-risk (LR) and HR-MSM in terms of number of partners and frequencies of sexual encounters. Total size of the population was 10,000 MSM, 3,000 of whom were categorized as HR-MSM.

The next paragraphs briefly summarize the main characteristics of the inter- and intra-host processes in the model for each scenario. In every scenario, ten cases of monkeypox were introduced among HR-MSM on day 1. All scenarios were run 100 times for 720 days. For further details, references and explanations for the assumptions made, please see Supplement 1.

### Baseline scenario

Scenario A was the baseline scenario to which the remaining scenarios were compared (Table 1). During each sexual encounter between an infectious and a susceptible individual, we assumed a 20% transmission probability of monkeypox. After a uniform incubation period of 7 days, exposed individuals became infectious for 21 days. Fifty per cent of infectious individuals were diagnosed with monkeypox after an average patient delay plus diagnostic delay of 14 days since the start of the infectious period. Diagnosed individuals ceased sexual activity for the next 28 days. The remaining undiagnosed individuals continued having sexual encounters. All cases recovered on day 21, after which lifelong immunity against reinfection was assumed.

**Table 1:** Model scenarios and results.

| Scenario | Probability of transmission per sexual encounter (%) | Proportion of undiagnosed cases (%) | Proportion of Contacts Traced (%) | PEP vaccination | PrEP vaccination = Proportion of HR-MSM vaccinated at day 1 (%) | Proportion of ongoing epidemics at day 720 (%) | Number of cases by day 720, median (IQR) | Epidemic duration, median (IQR)<sup>a</sup> | Number of people vaccinated, median (IQR) | Reduction in number of cases compared to scenario A (%) |
| --- | --- | --- | --- | --- | --- | --- | --- | --- | --- | --- |
| A | 20 | 50 | 0 | No | 0 | 55 | 1,442 (1,073 – 1,650) | 720 (621 - 720) | 0 | REF |
| B | 20 | 50 | 10 | No | 0 | 47 | 943 (636 – 1,284) | 690 (566 - 720) | 0 | 35 |
| C | 20 | 50 | 10 | Yes | 0 | 49 | 867 (591 – 1,168) | 714 (557 - 720) | 68 (46 - 82) | 40 |
| D | 20 | 50 | 10 | No | 1 | 43 | 924 (533 – 1,229) | 682 (558 - 720) | 30 (30 - 30) | 36 |
| E | 20 | 50 | 10 | No | 2.5 | 37 | 824 (493 – 1,044) | 631 (494 - 720) | 75 (75 - 75) | 43 |
| F | 20 | 50 | 10 | No | 5 | 29 | 632 (327 - 865) | 595 (409 - 720) | 150 (150 - 150) | 56 |
| G | 20 | 50 | 10 | No | 10 | 13 | 321 (188 - 525) | 408 (280 - 596) | 300 (300 - 300) | 78 |
| H | 20 | 50 | 10 | No | 25 | 0 | 136 (95 - 195) | 235 (171 - 314) | 750 (750 - 750) | 91 |
| I | 20 | 50 | 10 | No | 50 | 0 | 72 (57 - 86) | 131 (105 - 157) | 1,500 (1,500 – 1,500) | 95 |
| X | 10 | 50 | 0 | No | 0 | 0 | 71 (56 - 85) | 138 (114 - 190) | 0 | - |
| Y | 30 | 50 | 0 | No | 0 | 2 | 3,812 (3,660 – 3,932) | 532 (503 - 585) | 0 | - |
| Z | 20 | 0 | 0 | No | 0 | 0 | 185 (113 - 296) | 277 (198 - 404) | 0 | - |

<sup>a</sup> this number represents an underestimation as epidemics that were still ongoing at day 720 were assumed to last 720 days IQR = interquartile range; MSM = men who have sex with men; HR-MSM = high-risk MSM; PEP = post-exposure prophylactic (vaccination); PrEP = pre-exposure prophylactic (vaccination)

### Undiagnosed infections

To evaluate the impact of undiagnosed infections on the epidemic, scenario Z provided an alternative to scenario A in which 0% of infections remained undiagnosed.

### Per-encounter transmission probability

Scenarios X and Y were identical to scenario A, except for the per-encounter monkeypox transmissibility probability, which was set to 10% and 30%, respectively.

### Partner notification, post-exposure vaccination and pre-exposure vaccination

In scenarios B to I, individuals diagnosed with monkeypox notified 10% of their partners of the last 21 days prior to diagnosis. All notified partners ceased sexual activity for the next 28 days. Additionally, in scenario C, notified partners of the last seven days prior to the index partner’s diagnosis were vaccinated (post-exposure vaccination). In scenarios D to I, pre-exposure vaccination was done at day 1 of the model, in 1% to 50% of HR-MSM. Both pre- and post-exposure vaccination were assumed to prevent infection in 85% of vaccinees and have a lifelong effectiveness against infection. Childhood smallpox vaccination was not taken into account in the model.

### Sensitivity analysis

In a sensitivity analysis, we repeated all scenarios, while introducing one additional monkeypox case per week among HR-MSM, which represents an infection imported by travel.

## Results

The baseline scenario, in which half of the monkeypox cases remained undiagnosed, resulted in a median of 1,442 (IQR 1,073 - 1,650) cases by day 720 (Table 1 and Figure S1 in Supplement 2). This was almost eight-fold higher than scenario Z, in which all cases were diagnosed (median of 185, IQR 113 – 296 cases). Simulations with 10% and 30% transmission probability per sexual encounter resulted in unrealistically small (median 71, IQR 56 – 86 cases) or large (3,812, IQR 3,660 – 3,932 cases) epidemics, respectively.

If 10% of contacts of diagnosed cases abstained from sex (scenario B), the median number of cases by day 720 was reduced to a median of 943 (IQR 636 – 1,284), which represents a 35% reduction compared to baseline (Table 1 and Figure S2 in Supplement 2). Post-exposure vaccination of 10% of contacts (scenario C) had relatively limited additional impact (40% reduction compared to scenario A, to a median of 867, IQR 591 – 1,168 cases). It also required a median of 68 (IQR 46 – 82) contacts to be vaccinated and did not reduce epidemic duration compared to scenario B. Pre-exposure vaccination of a comparable number of HR-MSM (n = 75, scenario E) at day 1 was slightly more effective than post-exposure vaccination (reduction of 43% of cases compared to scenario A). Pre-exposure vaccination of 5%, 25% and 50% of HR-MSM resulted in a 56%, 91% and 95% reduction in number of cases, respectively. The epidemics in the sensitivity analysis were much larger and more protracted, with much lower impact of all interventions on epidemic size. None the less in this analysis, pre-exposure vaccination of 150 HR-MSM reduced the epidemic size to a greater extent than post-exposure vaccination of a similar number of contacts (Table S1 in Supplement 2).

## Discussion

The results of this model suggest that undiagnosed monkeypox infections may have an important impact on the epidemic. Secondly, our findings suggest that contact tracing helps to reduce epidemic size even if only 10% of contacts effectively ceased sexual activity. Finally, if only a small proportion of partners can be traced, post-exposure vaccination of those partners may be less effective than vaccinating a random proportion of individuals at highest risk of infection, and in our model this effect became more pronounced in scenarios with a weekly influx of new cases from other endemic/epidemic regions via travel.

The data presented here should be interpreted in the context of the design of the model and the assumptions made to parameterize it. We currently do not have accurate estimates of key parameters such as the proportion with unrecognized infections and the per-encounter transmission probability and how this varies according to type of (sexual) contact. In addition, our model did not capture superspreading events, which may have played an important role in the current outbreak. Finally, we modelled a relatively limited set of parameters.

Network-based models such as the one used here are particularly suitable to study transmission of an infectious disease in a densely connected sexual network of MSM. They have a proven utility in modelling other STIs such as gonorrhea and HIV, [11] and are likely to provide a more accurate representation of the sexual networks responsible for STI spread than the branching process models previously used to model monkeypox transmission among MSM. [6,8]

In conclusion, our model emphasizes the need to quantify key parameters such as transmission probability and the proportion of monkeypox infections that are unrecognized. A key finding is that pre-exposure vaccination of individuals at highest risk of infection may be more effective than post-exposure contact vaccination.

## Data Availability

The code used for the model is available from the corresponding author.

https://www.thelancet.com/journals/laninf/article/PIIS1473-3099(20)30778-7/fulltext

https://www.cambridge.org/core/journals/epidemiology-and-infection/article/current-levels-of-gonorrhoea-screening-in-msm-in-belgium-may-have-little-effect-on-prevalence-a-modelling-study/256A7F683A246E2B661AA89A73C062BA

## Notes

## Acknowledgements

Nil

## Potential conflicts of interest

None to declare. All the authors declare that they have no conflicts of interest

## Availability of data and materials

The code used for the model is available from the corresponding author

## Funding

This research did not receive any specific grant from funding agencies in the public, commercial, or not-for-profit sectors.

## Authors’ contributions

CK, AT and CVD conceptualized the study and, CVD and AT analyzed the data and drafted the manuscript, CK and NH revised the manuscript. All authors reviewed and approved the final manuscript.

## Notes

### Competing Interest Statement

The authors have declared no competing interest.

### Author Declarations

The study used ONLY openly available human data that were originally located at: https://www.cambridge.org/core/journals/epidemiology-and-infection/article/current-levels-of-gonorrhoea-screening-in-msm-in-belgium-may-have-little-effect-on-prevalence-a-modelling-study/256A7F683A246E2B661AA89A73C062BA https://www.thelancet.com/journals/laninf/article/PIIS1473-3099(20)30778-7/fulltext

