## Supplement 1 for "The roles of unrecognized monkeypox cases, contact isolation and vaccination in determining epidemic size in Belgium. A modelling study"

### **Supplement 1: Technical Appendix EpiModelMPX: Network model for simulating monkeypox transmission among MSM.**

### 1. **General description**

This document describes a network model to explain transmission of monkeypox (MPX) among men who have sex with men (MSM) in Belgium. The project is an extension of the EpiModel platform (<https://www.epimodel.org/>) and is based on the existing R package EpiModelHIV by Jenness et al (<https://github.com/EpiModel/EpiModelHIV>). Similar to EpiModelHIV, the current project uses the statistical framework of separable temporal exponential-family random graph models (STERGMs) to fit and simulate dynamic networks. The framework has been extensively described in Goodreau et al. [1] and Jenness et al. [2,3], as well as in the EpiModel page (www.epimodel.org). Statistical software R v.4.03 [4] and packages Epimodel [5] v2.0.3, tergm [6] v.3.7.0, tergmLite [7] v2.2.1 and ergm [8] v3.11.0 were used for the programming of this model.

### 2. **Data**

The parameters in the model come from two separate sources. Parameters regarding low-risk (LR-)MSM come from a previously published modelling study by Buyze et al., using the Belgium-based participants of the 2010 European MSM Internet Survey (EMIS). [9] The parameters and code used in this modelling study is open access. [9] Parameters for the high-risk (HR-)MSM were estimated using individual data from the PReGo study. [10] In the PReGo study, a cohort of 343 HR-MSM were followed up as part of a clinical trial which aimed to assess the efficacy of a mouthwash to prevent sexually transmitted infections (STIs). Data from both sources included information on the network consistency, partnership formation and dissolution, as well as behavioral and epidemiological characteristics.

Assumptions on Monkeypox natural history and transmission came from several research papers, as explained in detail below.

### 3. **Model structure**

The model consists of two parallel, interacting networks representing main and casual partnerships, in order to be able to preserve behavioral characteristics linked to each partnership type. The two parallel networks contained the same set of persons who were further distinguished into two STI risk groups (HR-MSM and LR-MSM) to account for behavioral patterns within the MSM population.

The formation of partnerships in the two networks (main and casual) was governed by similar formation equations. More specifically, the formation of main (casual) partnerships depended on the average number of partnerships currently in the network and the risk-group an individual belonged to. All those characteristics were targeted to match observed statistics in the study by Buyze et al. and the PreGo study, and were held constant over time. For both main and casual partnerships, there was a constant hazard of relationship dissolution depending on the relationship type. We initialized a population of 10,000 Belgian MSM, 30% of whom were randomly allocated to the high-risk group. [11,12]

### 3.1. **Partnership formation and dissolution**

The number of ongoing partnerships (main and casual) and their combination were derived from the available data for each of the two risk groups. We allowed each individual in the model to have either zero, one or more than one main partner and either zero, one or more than one casual partner, per time point. The distribution of main and casual partnerships by risk-group is presented in Table S1.

Table S1: Partnership formation probabilities stratified by partnership type and low/high risk group

|  |  | Casual partners | |
| --- | --- | --- | --- |
| Low-risk MSM | Main partners | 0 | > 0 |
|  | 0 | 42.46 % | 7.24 % |
|  | 1 | 38.98 % | 7.94 % |
|  | > 1 | 2.36 % | 1.01 % |
| High-risk MSM | Main partners |  |  |
|  | 0 | 12.16 % | 27.94 % |
|  | 1 | 11.80 % | 21.8 % |
|  | > 1 | 7.99 % | 18.32 % |

MSM = men who have sex with men

The formation of main and casual partnerships was affected by the total number of partnerships currently in the network and the risk-group of the individual. The partnership dissolution was modelled as a memoryless process with a single parameter depending on the relationship type. The duration of main partnerships was based on an approximation from a Dutch modelling study and was set at 1355 days. The duration of casual partnerships was set deterministically at 10 days. [13–15]

### 4. **Model processes & simulations**

Once the network of interest was initialized, the following steps occurred at each simulation:

At time step 1 (initialization)

1. Random allocation of infection status with MPX
2. Random allocation of vaccination status
3. Increment time by one step

At time step > 1:

1. Update of network in terms of creating new/dissolving current relationships.
2. Calculation of the number of sex acts that will occur in each active partnership.
3. Probabilistic transmission of MPX in case of discordant partnerships.
4. Start/end of incubation period
5. Start/end of infectious period
6. Cessation/resuming of sexual activity of diagnosed cases and/or their sexual contacts
7. Vaccination of sexual partners of diagnosed cases
8. Recovery from a MPX infection
9. (in sensitivity analysis 2: introduction of additional MPX cases)
10. Calculation of epidemiological statistics
11. Increment time by one step.

In this document as well as in the main text, a month refers to a period of 30 days, and a year is a period of 360 days. All scenarios were simulated 100 times for two years (720 days). The scenarios are described in the main paper (Table 1). The merged data from all 100 simulations were used to analyze each scenario.

#### 4.1. **Initialization**

4.1.1. **Seeding**

Ten monkeypox cases were randomly seeded among HR-MSM at the start of their incubation period.

4.1.2. **Population-based vaccination**

Depending on the scenario, 0 to 50% of HR-MSM were randomly selected for vaccination (see also section 4.3.1 and 4.4.2).

#### 4.2. **Inter-host processes**

In this section we present the processes of the model that occurred between two nodes in the network that shared interacted through a sex act on a given time step.

4.2.1. **Sex acts**

At each time step of the simulation, a list of active partnerships and their respective type (main or casual) was created based on the current composition of the network. Based on the type of relationship and the risk group combination of the partnership, occurrence of a sex act between the two partners on a given time step was calculated by random draws from a Bernoulli distribution with a mean value depending on the type of partnership and the risk-group of the two partners, as shown in Table S2. The mean sex act rate for mixed partnerships (a high- and a low-risk partner) was not available in our data sources and the average of the act rates of the two homogenous partnership combinations was assumed instead.

Table S2: Sex act rate probabilities per partnership type

|  | **Main partnerships** | | **Casual partnerships** | |
| --- | --- | --- | --- | --- |
|  | Low-risk | High-risk | Low-risk | High-risk |
| Low-risk | 0.2348208 |  | 1 |  |
| High-risk | 0.2348208 | 0.2348208 | 1 | 1 |

4.2.2. **Transmission**

Directional transmission of monkeypox occurred stochastically between discordant partners that engaged in a sex act. Depending on the scenario, the per-encounter transmission probability was assumed to be 10% to 30% (Table S3). This parameter represents the average probability that the virus is transmitted from one partner to another through the entire spectrum of sexual acts that may occur during one encounter. Transmission was modeled as a stochastic process based on a random draw from a Bernoulli distribution.

#### 4.3. **Intra-host processes**

4.3.1. **SEIARV model**

We used a SEIARV model (Table S3, Figure S1). Susceptible individuals (S) were assumed to go through a 7-day incubation period (E) during which they are not infectious, and to become infectious (I or A) for the next 21 days. Depending on the scenario, 50 to 100% of infected individuals would eventually start noticing symptoms, get diagnosed, cease sexual activity, and/or notify their contacts. Whether an infected individual were to follow this path of events (I) or remain sexually active throughout his entire infectious period (A) was determined at the time of transmission by a random draw from a Bernoulli distribution. For individuals with status I, the time step after the incubation period at which their diagnosis would be made was determined by a random draw from a normal distribution with mean 14 days.

Figure S1: SEIARV model

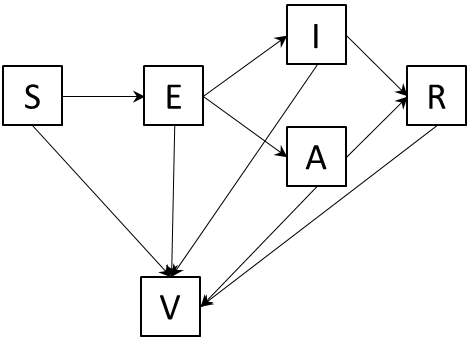

S = susceptible; E = exposed; I = infectious and eventually gets diagnosed; A = infectious but remains undiagnosed; R = recovered; V = vaccinated

Table S3: Monkeypox-related parameters and references

| **Status** | **Parameter** | **Value** | **Explanation** | **Reference** |
| --- | --- | --- | --- | --- |
| S | Susceptible period | Until infected |  |  |
| ß | Per-encounter transmission probability | 10% to 30% | One meta-analysis of studies in the context of Congo Basin clade outbreaks of monkeypox estimated a secondary attack rate among unvaccinated household contacts of about 10%. Data for the West-African clade are lacking. We assumed that the average (per-encounter) sexual-associated transmission probability is higher than the average within-household (cumulative) transmission probability. | [16] |
| E | Incubation period (non-infectious) | 7 days (uniform distribution) | Estimated at 5-21 days. | [17–19] |
| I | Infectious period (individuals who would eventually get diagnosed) | 21 days (uniform distribution) | Individuals with status “I” ceased sex after diagnostic delay δ | [17,19] |
| δ | Diagnostic delay in individuals with status “I” | 14 days (normal distribution around day 14) | Expert opinion (pre-symptomatic period + early symptomatic period + symptomatic period during which the infected individual continues having sexual encounters) |  |
| A | Infectious period (individuals who remain undiagnosed) | 21 days (uniform distribution) | Individuals with status “A” continued having sexual encounters throughout the entire infectious period, because their infection remained undiagnosed, failure to comply with the recommendation to abstain from sex, etc. | [17,19] |
| R | Recovery period (immunity against reinfection) | Lifelong | Mathematical modelling studies of smallpox have assumed lifelong immunity after infection. | [17] |
| V | Vaccinated immunity against infection | Lifelong | Mathematical modelling studies of smallpox have assumed lifelong immunity after vaccination. | [17] |
| є | Vaccine efficacy (in the context of pre- or post-exposure prophylaxis) | 85% | Studies in the context of Congo Basin clade outbreaks of monkeypox estimated 85% efficacy of childhood smallpox vaccination against monkeypox disease in close household contacts.  Data on vaccine effectiveness for post-exposure prophylaxis are lacking. According to expert opinion, older generation smallpox vaccines were about 80% effective in preventing smallpox disease if given within 3 days after exposure. | [19–21] |

Individuals who ceased sexual activity did so for 28 days (= sum of incubation period + infectious period). A period of ceased sexual activity could overlap with any status of the SEIARV model.

Infected individuals (I or A) recovered from their infection with lifelong immunity against reinfection (R).

Depending on the scenario, vaccination could be done at day 1 (population-based pre-exposure vaccination) or later on in the model (partner-based post-exposure vaccination). Vaccinated individuals were assumed to have an 85% chance to be protected against infection, for the rest of their lives. In the model, for every individual eligible for vaccination (selected among S, E, I, A or R), a random draw from a Bernoulli distribution determined which individuals would be effectively immune by vaccination. These individuals received status V. Those not effectively protected by the vaccine kept their status as if they were not vaccinated.

#### 4.4. **Interventions**

4.4.1. **Partner tracing**

Depending on the scenario, 0% or 10% of sex partners in the last 21 days prior to diagnosis of diagnosed individuals (see section 4.3.1) were traced. An infected individual’s traceable partners (with status S, E, I, A, R or V) were identified among all his partners by a random draw from a Bernoulli distribution. All traceable partners ceased sexual activity for 28 days starting from the time step at which their infected index partner was diagnosed. As such, traced partners in our model are to be interpreted as those partners who are both traceable and able to interrupt further onward transmission of the virus. This 10% level was based on the experience in our and other Belgian STI clinics that tracing more than an average of 10% of partners of HR-MSM is generally not feasible.

4.4.2. **Vaccination**

A) Population-based pre-exposure vaccination

Depending on the scenario, 0 to 50% of HR-MSM were vaccinated at day 1.

B) Partner-based post-exposure vaccination

Depending on the scenario, 0 to 100% of traced partners got vaccinated if their most recent sex act with the infected index partner found place in the last seven days prior to diagnosis of their index partner.

#### 4.5. **Sensitivity analysis 2**

Sensitivity analysis 2 took into account the introduction of new monkeypox cases that were imported through interaction of HR-MSM with networks outside of the model (e.g. a HR-MSM who returned from a trip abroad where he was exposed to monkeypox. In the model this was simulated by the weekly allocation of status “E” to one random HR-MSM who previously had status “S”.

### 5. **Bibliography**

6.	Krivitsky PN, Handcock MS. tergm: Fit, Simulate and Diagnose Models for Network Evolution Based on Exponential-Family Random Graph Models. 2020.

7.	Jenness SM, Klumb C. tergmLite: Fast Simulation of Simple Temporal Exponential Random Graph Models. 2020;

8.	Hunter DR, Handcock MS, Butts CT, Goodreau SM, Morris M. ergm: A Package to Fit, Simulate and Diagnose Exponential-Family Models for Networks. J Stat Softw **2008**; 24:nihpa54860.

9.	Buyze J, Vanden Berghe W, Hens N, Kenyon C. Current levels of gonorrhoea screening in MSM in Belgium may have little effect on prevalence: a modelling study. Epidemiol Infect **2018**; 146:333–338.

10.	Van Dijck C, Tsoumanis A, Rotsaert A, et al. Antibacterial mouthwash to prevent sexually transmitted infections in men who have sex with men taking HIV pre-exposure prophylaxis (PReGo): a randomised, placebo-controlled, crossover trial. Lancet Infect Dis **2021**; 21:657–667.

11.	Sherriff NS, Jones AM, Mirandola M, et al. Factors related to condomless anal intercourse between men who have sex with men: Results from a European bio-behavioural survey. J Public Heal (United Kingdom) **2020**; 42:1–13.

12.	Hansson D, Strömdahl S, Leung KY, Britton T. Introducing pre-exposure prophylaxis to prevent HIV acquisition among men who have sex with men in Sweden: Insights from a mathematical pair formation model. 2020.
