## Supplement 2 for "The roles of unrecognized monkeypox cases, contact isolation and vaccination in determining epidemic size in Belgium. A modelling study"

***Supplement 2: Supplementary tables and figures***

### Figure S1: Epidemic curves, final epidemic size and epidemic duration (scenarios A, X, Y, Z)

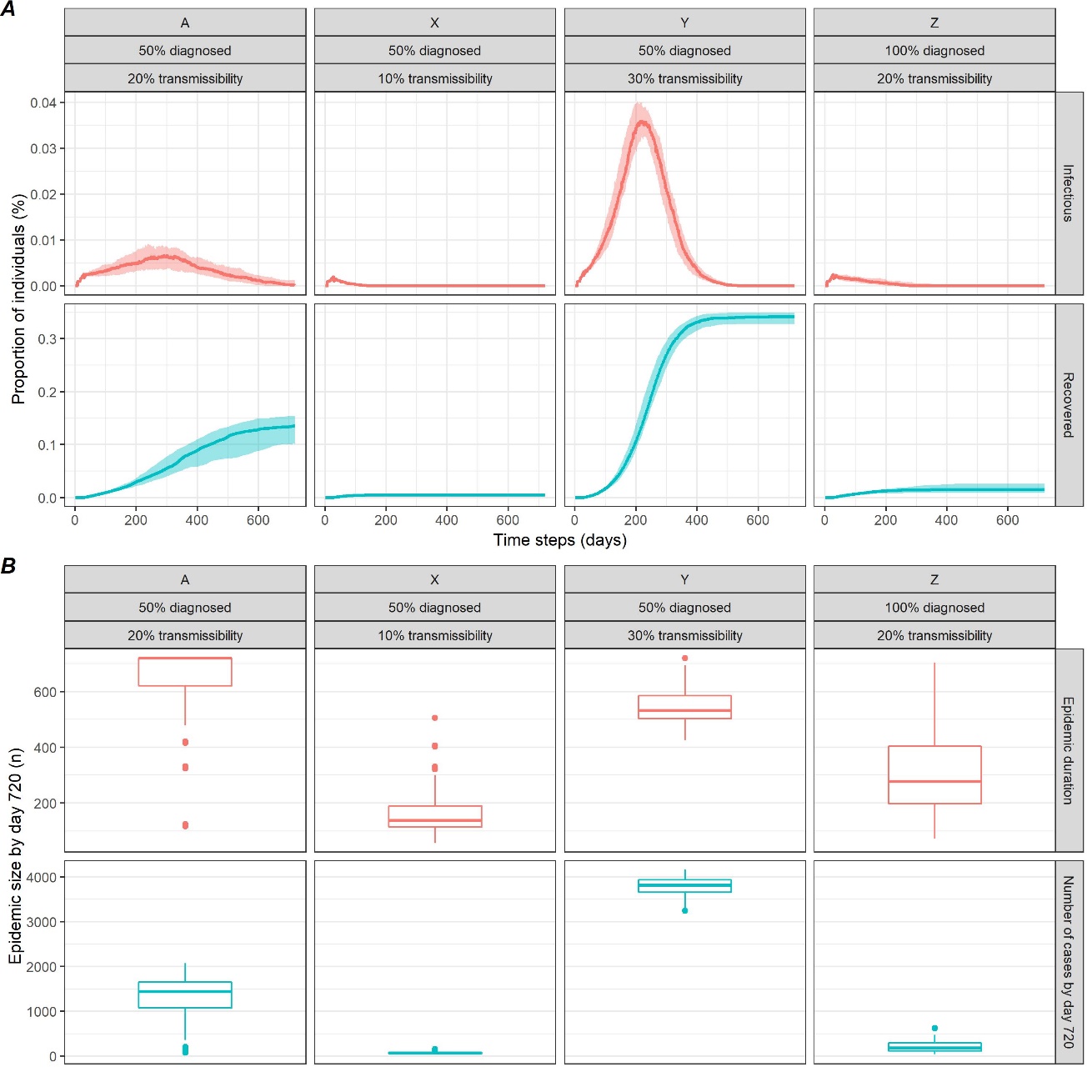

Figure S2: Epidemic curves, final epidemic size and epidemic duration (scenarios A to I)

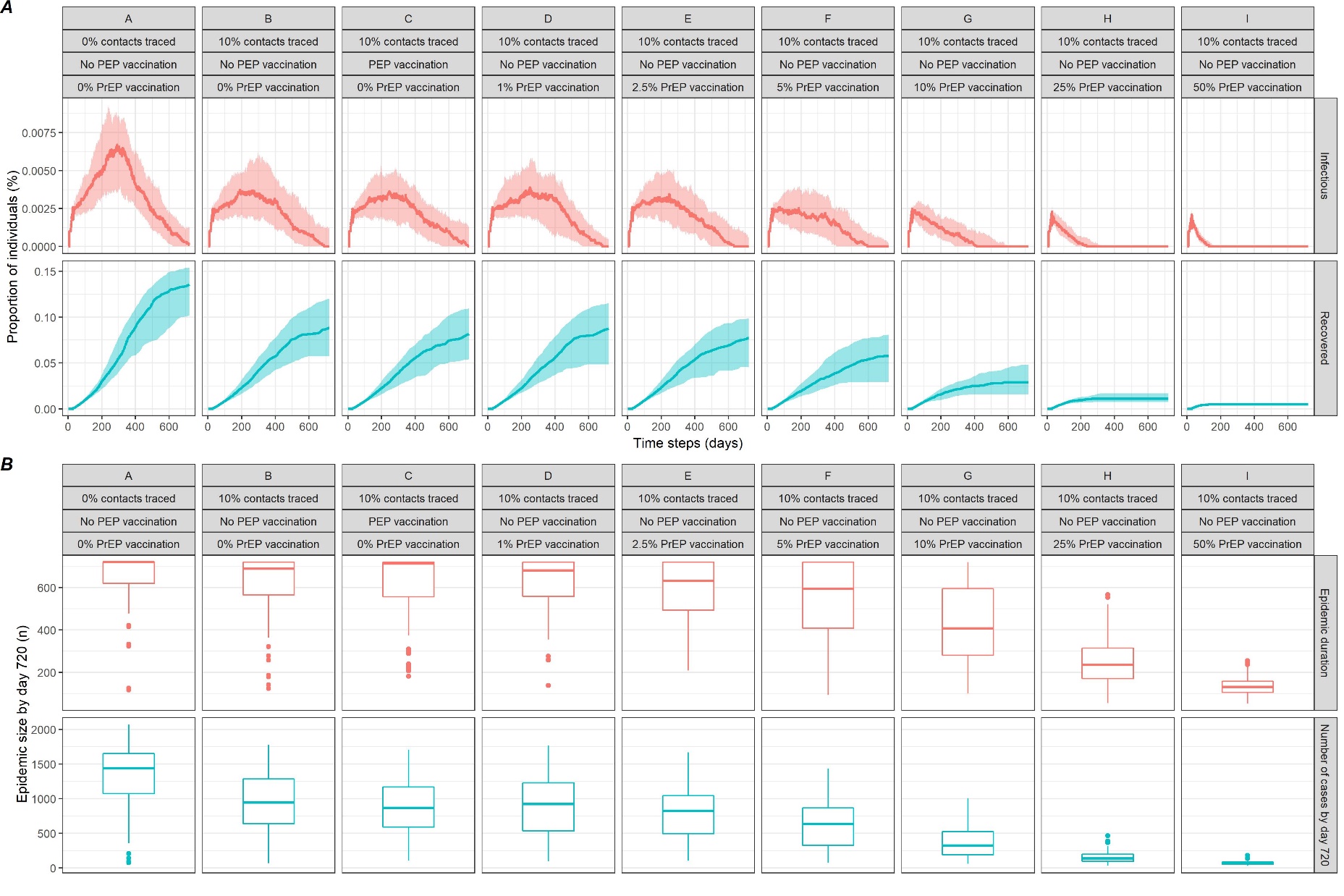

### Table S1: Sensitivity analysis - scenarios including weekly introduction of 1 additional monkeypox case among high-risk MSM

| **Scenario** | **Probability of transmission per sexual encounter (%)** | **Proportion of undiagnosed cases (%)** | **Proportion of Contacts Traced (%)** | **PEP vaccination** | **PrEP vaccination = Proportion of HR-MSM vaccinated at day 1 (%)** | **Proportion of ongoing epidemics at day 720 (%)** | **Number of cases by day 720, median (IQR)** | **Number of people vaccinated, median (IQR)** | **Reduction in number of cases compared to scenario A (%)** |
| --- | --- | --- | --- | --- | --- | --- | --- | --- | --- |
| A | 20 | 50 | 0 | No | 0 | 100 | 2,253 (2,114 - 2437) | 0 | REF |
| B | 20 | 50 | 10 | No | 0 | 100 | 1,969 (1,832 - 2,114) | 0 | 13 |
| C | 20 | 50 | 10 | Yes | 0 | 100 | 1,890 (1,750 - 1,980) | 140 (131 - 152) | 16 |
| D | 20 | 50 | 10 | No | 1 | 100 | 1,896 (1,760 - 2,042) | 30.6 (31 - 31) | 16 |
| E | 20 | 50 | 10 | No | 2.5 | 100 | 1,844 (1,752 - 1,950) | 75.3 (75 - 75) | 18 |
| F | 20 | 50 | 10 | No | 5 | 100 | 1,706 (1,554 - 1,819) | 150.6 (151 - 151) | 24 |
| G | 20 | 50 | 10 | No | 10 | 100 | 1,454 (1,338 - 1,540) | 300 (300 - 300) | 35 |
| H | 20 | 50 | 10 | No | 25 | 100 | 948 (886 - 1,005) | 750.6 (751 - 751) | 58 |
| I | 20 | 50 | 10 | No | 50 | 100 | 525 (493 - 559) | 1,500 (1,500 - 1,500) | 77 |
| X | 10 | 50 | 0 | No | 0 | 100 | 493 (469 - 529.5) | 0 | - |
| Y | 30 | 50 | 0 | No | 0 | 100 | 4,188 (4,062 - 4,306) | 0 | - |
| Z | 20 | 0 | 0 | No | 0 | 100 | 1,177 (1,068 - 1,267) | 0 | - |

IQR = interquartile range; MSM = men who have sex with men; HR-MSM = high-risk MSM; PEP = post-exposure prophylactic (vaccination); PrEP = pre-exposure prophylactic (vaccination)
